# Seizure likelihood varies with day-to-day variations in sleep duration in patients with refractory focal epilepsy: A longitudinal EEG investigation

**DOI:** 10.1101/2021.05.03.21256436

**Authors:** Katrina L. Dell, Daniel E. Payne, Vaclav Kremen, Matias I. Maturana, Vaclav Gerla, Petr Nejedly, Gregory A. Worrell, Lhotska Lenka, Filip Mivalt, Raymond C. Boston, Benjamin H. Brinkmann, Wendyl D’Souza, Anthony N. Burkitt, David B. Grayden, Levin Kuhlmann, Dean R. Freestone, Mark J. Cook

## Abstract

**Background:** While the effects of prolonged sleep deprivation (≥24 hours) on seizure occurrence has been thoroughly explored, little is known about the effects of day-to-day variations in the duration and quality of sleep on seizure probability. A better understanding of the interaction between sleep and seizures may help to improve seizure management.

**Methods:** To explore how sleep and epileptic seizures are associated, we analysed continuous intracranial EEG recordings collected from 10 patients with refractory focal epilepsy undergoing ordinary life activities. A total of 4340 days of sleep-wake data were analysed (average 434 days per patient). EEG data were sleep scored using a semi-automated machine learning approach into wake, stages one, two, and three non-rapid eye movement sleep, and rapid eye movement sleep categories.

**Findings:** Seizure probability changes with day-to-day variations in sleep duration. Logistic regression models revealed that an increase in sleep duration, by 1·66 ± 0·52 hours, lowered the odds of seizure by 27% in the following 48 hours. Following a seizure, patients slept for longer durations and if a seizure occurred during sleep, then sleep quality was also reduced with increased time spent aroused from sleep and reduced REM sleep.

**Interpretation:** Our results demonstrate that day-to-day deviations from regular sleep duration correlates with changes in seizure probability. Sleeping longer, by 1·66 ± 0·52 hours, may offer protective effects for patients with refractory focal epilepsy, reducing seizure risk. Furthermore, the occurrence of a seizure may disrupt sleep patterns by elongating sleep and, if the seizure occurs during sleep, reducing its quality.

**Funding:** Australian National Health and Medical Research Council, US National Institutes of Health and Czech Technical University in Prague and Epilepsy Foundation of America Innovation Institute

## Introduction

Sleep and epileptic seizures share a complex and bidirectional relationship.^1^ Deviations from normal sleep duration and quality can greatly influence the risk of a seizure.^2^ In turn, the occurrence and treatment of seizures can disrupt normal sleep patterns.^3-5^ Understanding the complexities of this relationship is an important step toward improving seizure management.

While it is generally accepted that total sleep deprivation for periods of 24 hours or longer can lead to seizures, even in individuals that do not have epilepsy,^6,7^ the role of partial sleep deprivation in promoting seizures remains controversial.^8,9^ Of four studies using self-reported sleep and seizure information, three indicated that partial sleep deprivation increased the probability of a seizure,^9-11^ while the fourth reported a small degree (< 1 hour) of sleep loss did not influence seizure occurrence at all.^8^ To our knowledge, no electroencephalography (EEG) investigations have addressed the role of partial sleep deprivation.

Similar uncertainty exists regarding the effects of sleep quality on seizure propensity in people with epilepsy. Typically, sleep quality is quantified by the absolute or percentage of time spent in each sleep stage as well as the degree of sleep fragmentation or time spent awake after sleep onset.^12^ Sleep with more frequent/longer arousals, reduced rapid eye movement (REM) sleep, or reduced stage three non-rapid eye movement (NREM) sleep is generally considered to be less restorative.^13^ Studies that have selectively manipulated REM sleep duration suggest that increased REM sleep may act to reduce cortical excitability and, in turn, seizure occurrence.^14,15^ However, it is yet to be determined if day-to-day variations in REM sleep, deep sleep, or sleep fragmentation can influence seizure occurrence in humans.

The influence of sleep on seizure risk is further complicated by the effects that seizures themselves have on sleep duration and quality. Short-term EEG investigations have shown following a focal impaired awareness or focal to bilateral tonic-clonic seizure, there is a reduction in the proportion of REM sleep. When the seizure occurs during sleep, this reduction in REM sleep is more pronounced and is accompanied by an increase in stage one NREM and decreases in stage two NREM and deep sleep.^5^ Similar disruptions have been observed for up to four nights following generalised convulsive status epilepticus, where sleep consists predominantly of stage one NREM with minimal REM and deep sleep.^16^ These alterations in sleep may play an important role in the generation of subsequent seizures.

Until now, long-term (>1-2 weeks) investigations of sleep and epilepsy have relied on seizure diaries. This is problematic as seizure diaries are highly unreliable, with patients often reporting less than 50% of their seizures.^17,18^ Furthermore, short-term EEG investigations do not have the statistical power to investigate the effects of subtle changes in sleep duration or quality on seizure occurrence. The small number of seizures collected per patient in a typical EEG study usually requires the data to be collapsed across patients for analyses, which is problematic given the high degree of variability that exists between individuals.^19,20^ Furthermore, the majority of EEG data is collected in a hospital setting, often with medication withdrawal and deliberate sleep deprivation, which is unlikely to be representative of the true sleep and seizure trends that occur in day-to-day life.

Here, we present the first study using long-term ambulatory intracranial EEG data (spanning several months to years for each patient) to investigate the relationship between sleep and seizures. Recordings were classified into sleep stages and analysed to describe sleep-wake patterns in our patient group. Our objective was to explore and define the bidirectional relationship between seizures and sleep duration and composition. Our results demonstrate that an increase in sleep duration is correlated with reduced seizure odds in the following 48 hours, which may imply that increased sleep duration offers protective effects. Variability in sleep stage proportions produced variable effects across patients, with no significant change in the odds of a seizure in the following 48 hours. The occurrence of a seizure is followed by longer sleep durations and, if a seizure occurs during sleep, sleep quality is reduced with a lower proportion of REM sleep and a greater proportion of time spent in brief arousal.

## Materials and Methods

### Data

The data used in this study were part of the first-in-man clinical trial of an implantable seizure advisory system which was approved by the Human Research Ethics Committees of the three participating clinical centres (LRR145/13).^17^ Patient selection prioritised suitable seizure frequencies (between 2 and 12 per month) and adults with sufficient independence to make the implanted seizure advisory device useful for managing daily activities. All patients gave written informed consent before participation in the clinical trial. Fifteen patients with focal epilepsy were implanted with an intracranial EEG device recording at 400 Hz. Periods where the external device was not in range of the transmitter or periods where the device was not charged caused dropouts in the data. The patient cohort showed a range of demographic and clinical features that is typical of the wider refractory epilepsy population, with a range of aetiologies, and including patients on multiple anti-seizure drugs, as well as patients on minimal therapy. The patient details and recording durations are provided in Table 1. Patients 3, 4, 5, 7, and 14 from the original trial were excluded from analyses as we did not have sleep information for these patients. There were no clear differences in the demographic or clinical features between included and excluded patient groups and thus we do not expect the exclusion of five to bias the results of this investigation. Sleep related comorbidities were ruled out for our patient group by clinical history and in-patient video EEG. For further details regarding the recruitment criteria, patient demographic and clinical procedures, see Cook et al. 2013.

**Table 1.**
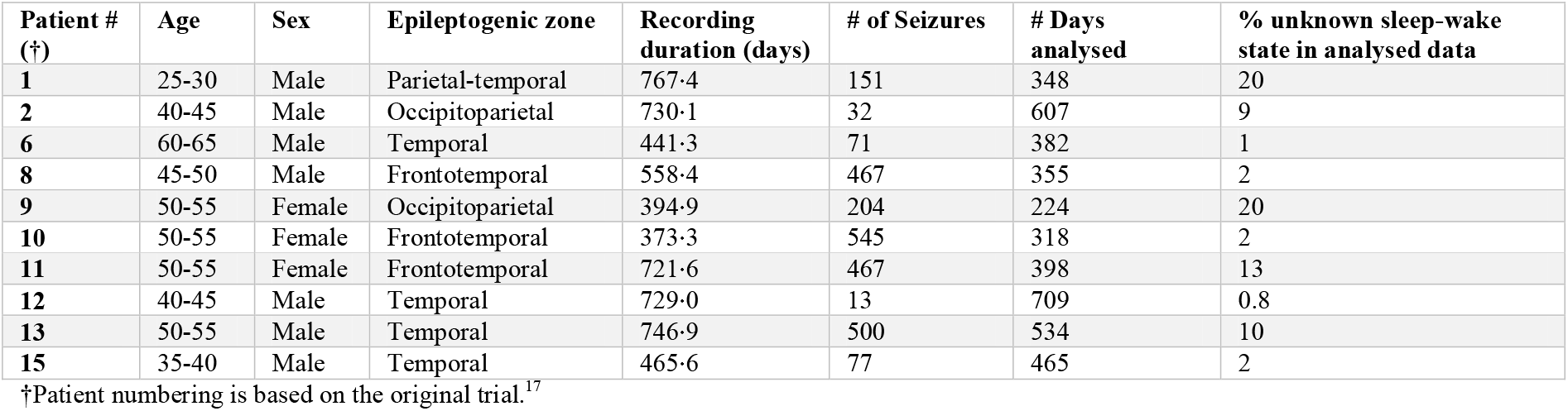
Patient demographics.

| Patient #<br>(†) | Age | Sex | Epileptogenic zone | Recording<br>duration (days) | # of Seizures | # Days<br>analysed | % unknown sleep-wake<br>state in analysed data |
| --- | --- | --- | --- | --- | --- | --- | --- |
| 1 | 25-30 | Male | Parietal-temporal | 767·4 | 151 | 348 | 20 |
| 2 | 40-45 | Male | Occipitoparietal | 730·1 | 32 | 607 | 9 |
| 6 | 60-65 | Male | Temporal | 441·3 | 71 | 382 | 1 |
| 8 | 45-50 | Male | Frontotemporal | 558·4 | 467 | 355 | 2 |
| 9 | 50-55 | Female | Occipitoparietal | 394·9 | 204 | 224 | 20 |
| 10 | 50-55 | Female | Frontotemporal | 373·3 | 545 | 318 | 2 |
| 11 | 50-55 | Female | Frontotemporal | 721·6 | 467 | 398 | 13 |
| 12 | 40-45 | Male | Temporal | 729·0 | 13 | 709 | 0.8 |
| 13 | 50-55 | Male | Temporal | 746·9 | 500 | 534 | 10 |
| 15 | 35-40 | Male | Temporal | 465·6 | 77 | 465 | 2 |

†Patient numbering is based on the original trial.<sup>17</sup>

Seizure detections were verified by certified clinicians and expert investigators with the aid of seizure diaries and audio recorded from the portable seizure advisory device.

### Sleep-wake scoring

Intracranial EEG was scored in 30-second epochs into awake, stage one NREM (NREM1), stage two NREM (NREM2), stage three NREM (NREM3), or REM sleep categories based on methods adapted from Kremen, Brinkmann^22^. These methods have previously been confirmed and validated on intracranial EEG data with concurrent polysomnography and gold standard sleep scoring according to AASM2012 rules across multiple investigations and have yielded an average accuracy of 94% with Cohen’s kappa of 0.8732 for NREM2, NREM3 and awake states, and with REM sleep included an average of 91% in humans and 94% in dogs (not shown). The accuracy of stage 1 NREM sleep was less reliable, presumably because it is relatively infrequent, accounting for very little of the total sleep duration, and is very similar electrographically to the awake state. For this reason, we do not draw any major conclusions regarding this sleep type. Furthermore, any spontaneous NREM1 occurring separate from other sleep scores were ignored when marking sleep onset and offset transitions and were not included in the total sleep duration.

Sleep scoring was conducted using one representative electrode or the median of all electrodes selected manually by reviewer judgement. For each patient, nine days at equidistant positions throughout the dataset were manually sleep-wake scored and used to train a patient-specific automated classifier. The data was classified using a K nearest-neighbours algorithm with K=3-50 clusters or a Naïve Bayes classifier. Each classifier used an optimized subset of 8 features selected for the individual patient by a sequential feature selection method from a set of 21 extracted features. The trained classifier scored 24-hour sections at a time.

A generalized deep learning convolutional feed-forward neural network (CNN) algorithm tested and published in Nejedly, Kremen^21,23^ was utilized to scan the data for artifactual segments. The input signal was processed by the generalized CNN model and the resulting artifact probability matrix was used to analyse and interpret the artifactual segments of the data. The sleep-wake scored data was then pruned by removing any artefactual epochs or epochs/days with more than 50% of data missing. Unusable epochs/days were classified as ‘unknown’.

### Feature extraction

We used a subset of algorithms previously defined by Gerla, Kremen^24^ to extract spectral, entropy-based and wavelet transform features. We used 21 features extracted for each epoch, for each patient. Features included mean dominant and spectral median frequencies, mean absolute and mean relative spectral powers in the following frequency bands: 1-3 Hz (delta), 3-7 Hz (theta), 7-12 Hz (alpha), 12-15 Hz (low beta), and 15-20 Hz (high beta). As well as a spectral entropy feature, which is an estimate of the probability density function for each epoch of EEG signal in the frequency bands described above and for an additional 1-25 Hz band. To overcome energy fluctuations in the data, individual features were normalised into scale given by 0.05 and 0.95 quantiles of each feature distribution. Classifiers were trained with normalised and unnormalized features and we selected the classifier that produced the best results for each patient. An example of the standard frequency band features together with a continuous wavelet scalogram and sleep scoring is shown in Figure 1.

**Figure 1.**
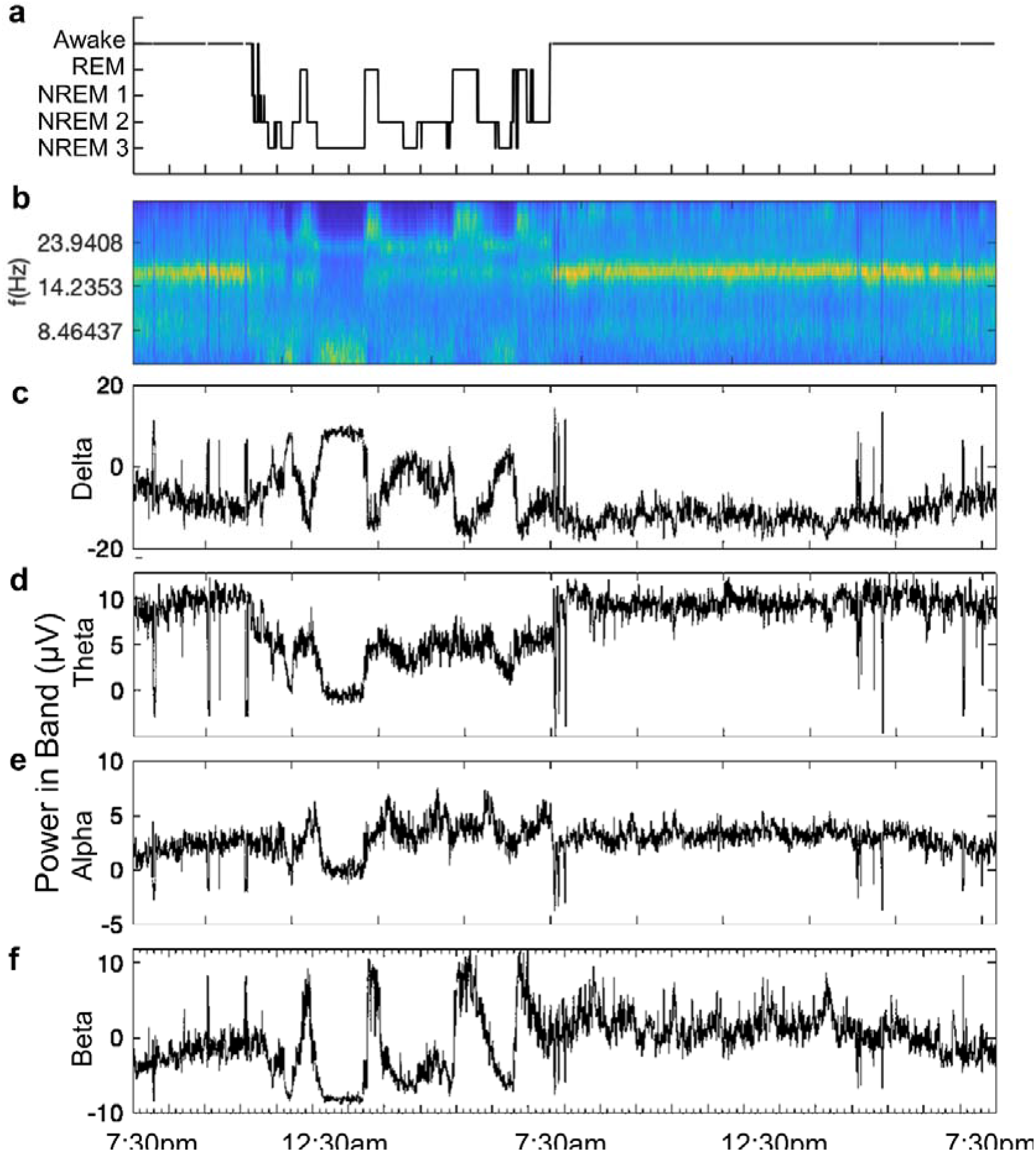
Power in Band features with continuous wavelet scalogram. A) Automated sleep scores and corresponding B) EEG time series continuous wavelet scalogram and power (μV) in Berger bands; C) delta (1-4 Hz), D) theta (4-7 Hz), E) alpha (8-12 Hz), and F) beta (12-30 Hz). Here, we show a recording from patient 15 on day 19 into the study.

### Statistical analysis

All sleep-wake patterns and the influence of seizures on sleep duration and structure were analysed using repeated measures one-way ANOVA. Bounded data were arcsine transformed prior to ANOVA. We did not assume sphericity of the data and so a Greenhouse-Geisser correction was used for all repeated measures ANOVA. To be consistent with existing literature, the interaction between seizure risk and day-to-day variability in sleep duration and composition were analysed using mixed effects logistic regression models. A p-value of 0.05 or lower was considered statistically significant unless otherwise stated. Data analyses were conducted using MATLAB R2017a, ANOVA was conducted using Graphpad Prism 8.3 and logistic regression modelling was conducted using Stata 16.1.

### Role of the funding source

The funders had no role in study design, data collection, analysis, interpretation, or the writing of the report. The corresponding author had full access to the data and had the final responsibility for the decision to submit for publication.

## Results

### Overview of sleep and seizure timing

A total of 4340 days of sleep-wake data were analysed across 10 patients (between 224 and 709 days per patient). As sleep was commonly interrupted by brief arousals, the main transitions into (“sleep onset”) and out of (“sleep offset”) sleep were manually labelled by visual inspection (Figure 2A). Any sleep sections shorter than 2 hours in duration were classed as naps. Previous reports by our group have described circadian and circaseptan patterns in seizure occurrence for this patient cohort.^25^ In Figure 2B we show the distributions of sleep onset, offset and seizure times across a 24-hour period. Of note, for patients 8 and 10 the seizures cluster at the same times of day as the regular sleep onset and offset transitions. This suggests that the timing of sleep could play an important role in seizure precipitation, at least in some patients. We also explored the circaseptan period but did not observe any clear trends between seizure occurrence and sleep transition times (Supplementary Figure 1).

**Figure 2.**
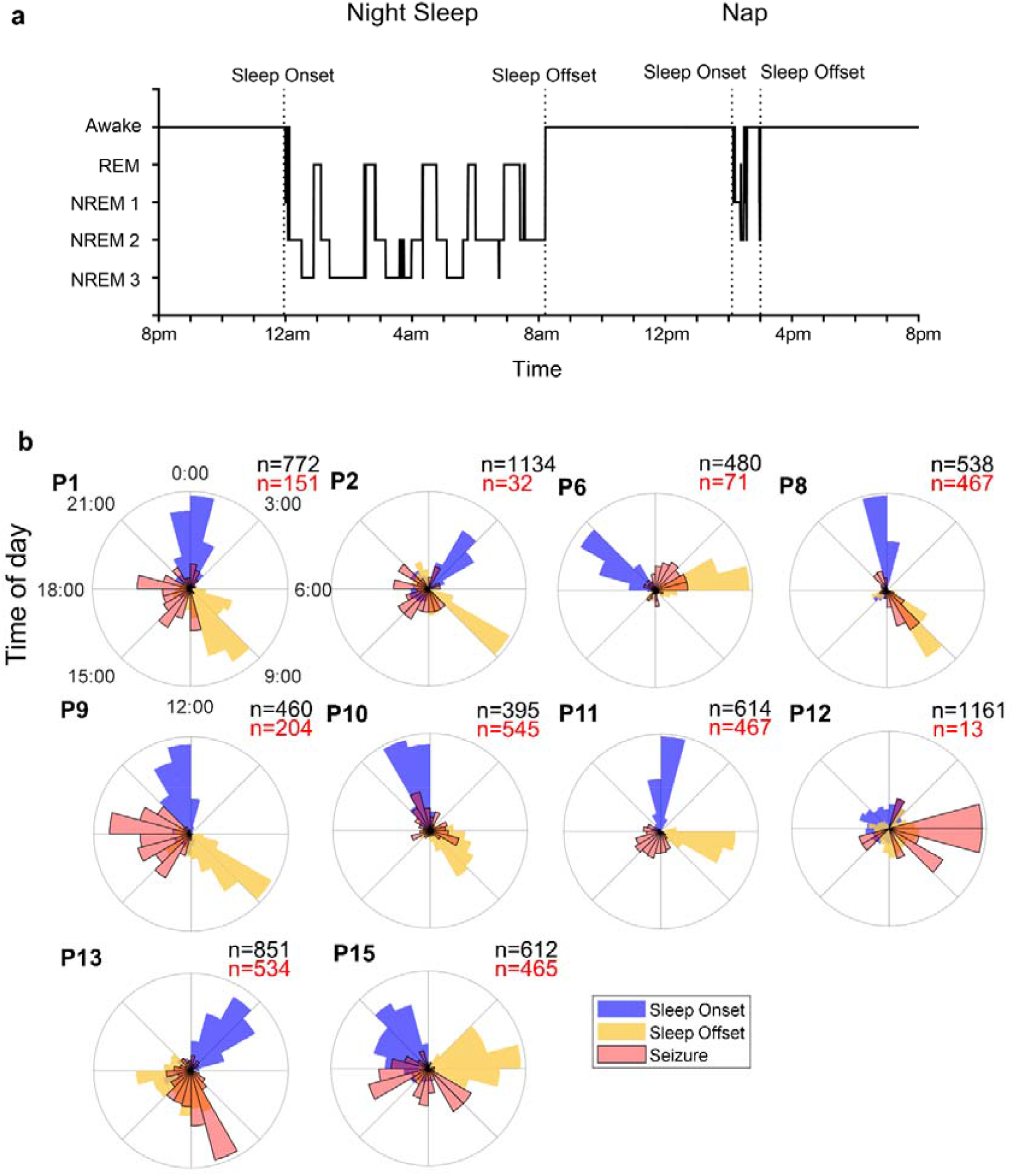
Sleep and seizure circadian cycles. A) Sleep-wake scores with example sleep transitions (sleep onset and sleep offset) labelled by visual inspection and indicated by dotted vertical lines. Sections of sleep shorter than 2 hours in duration were classified as naps. B) Normalized polar histograms of sleep onset (blue), sleep offset (yellow) and seizure (red) times throughout a 24-hour period. For each patient the total number of sleep transitions (onset or offset) is shown in black and the total number of seizures in red.

### Overview of sleep structure

Proportions of time in each sleep-wake state are shown for each patient in Figure 3A. The time spent in brief arousal from sleep was labelled as wake after sleep onset (WASO) and is included in the total sleep duration. Segments where sleep-wake scoring was not possible due to noise, artefacts, or data dropout are represented as ‘unknown’. Days that contained more than 30% unknown were excluded from all analyses. The average hours asleep for each patient ranged from 7.52 to 11.35 hours per seizure-free day including naps (Figure 3B). This is 0.21 to 4.04 hours longer than the average sleep duration reported for Australian adults, 7.31 hours.^26^ The longer duration of time spent asleep in our cohort may be the result of frequent napping, with patients taking an average of 0.21-1.04 naps on seizure-free days and 0.32-1.34 naps on days that seizures occurred (Figure 3C).

**Figure 3.**
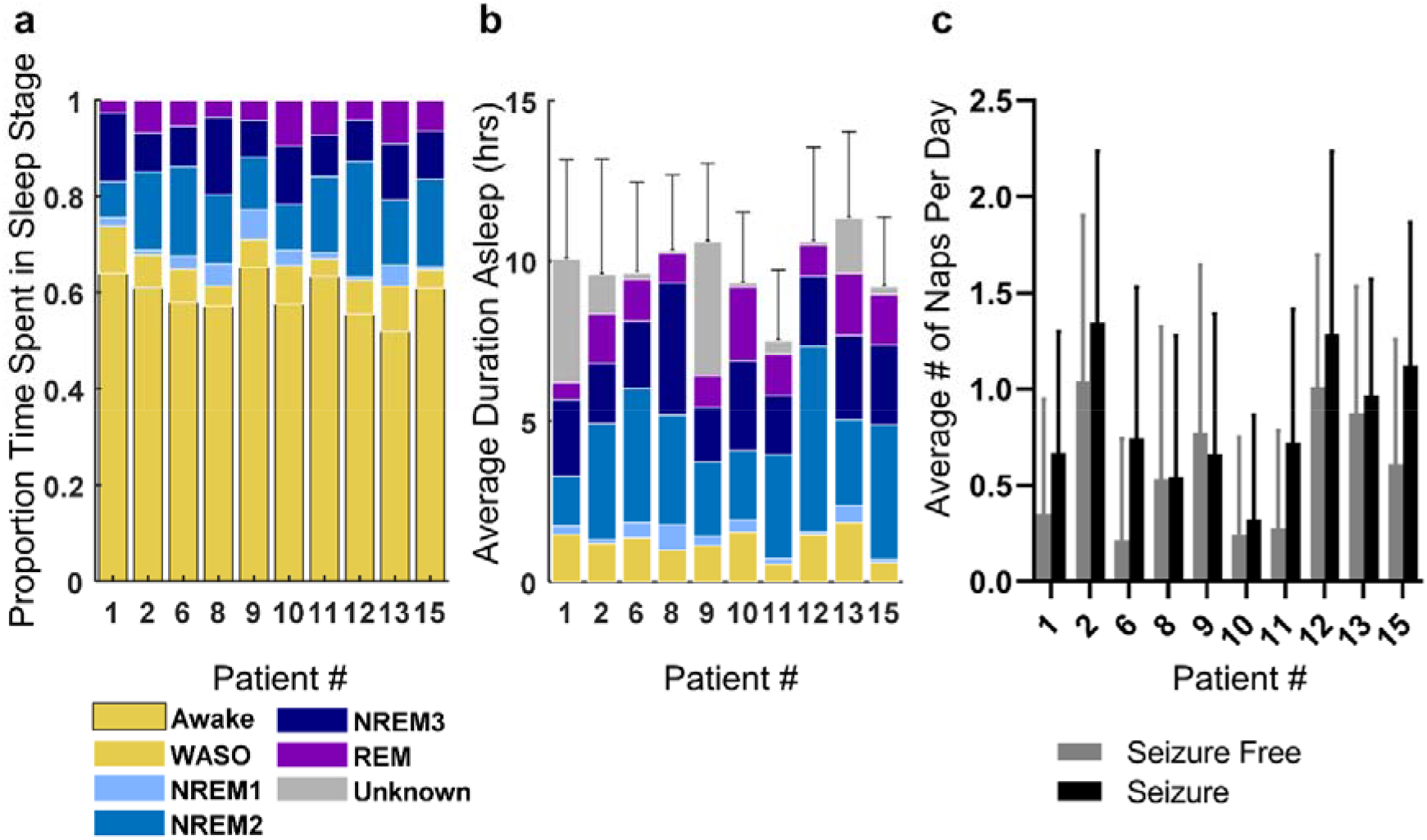
Sleep duration and composition overview. A) The proportion of time spent in each sleep-wake state, B) the average daily duration (hrs) spent asleep on seizure free days (night sleep + naps), and C) the average number of naps per seizure free and seizure days are shown for each patient. Error bars represent standard deviation

In order to determine if the patients’ sleep followed a typical architecture, sleep was divided into cycles. Cycle one was defined to begin at the first instance of REM sleep and continue until the next instance of REM sleep. Each subsequent cycle began at the last REM period and extended until the next, as illustrated in Figure 4A. Only the first five cycles of sleep, excluding naps, on seizure-free days are included in these analyses. Throughout sleep, cycle duration decreased with each subsequent cycle (Figure 4B). The time spent in NREM1 and REM sleep increased with each cycle while the time spent in NREM3 sleep decreased (Figure 4C). The trends were consistent across patients and the overall structure of sleep followed trends similar to those observed in healthy individuals.^27^ These results thus indicate our sleep scoring methods were effective.

**Figure 4.**
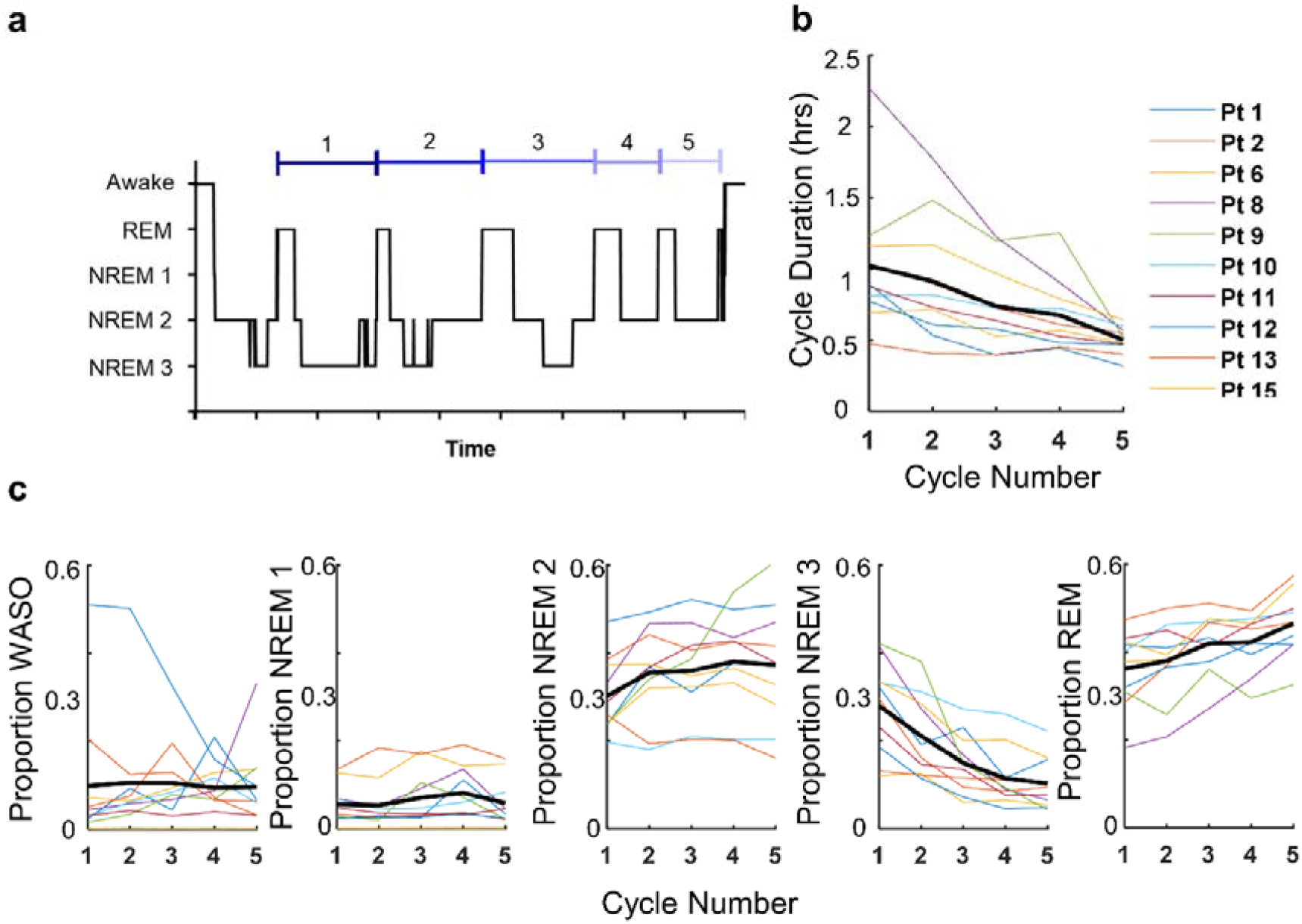
Night sleep structure. A) Example illustration of sleep cycles. The average B) cycle duration (hrs) and C) proportion of the cycle spent in each sleep stage is shown for each patient in colour with the population average represented by the bold black line.

### The relationship between sleep-wake state and seizure occurrence and duration

To explore how seizure characteristics related to current sleep-wake state on seizure, we calculated the proportion, rate, and duration of seizures in each sleep-wake category. Two patients (8 & 15) experienced more seizures during the wake state while the remaining eight patients (1, 2, 6, 9, 10, 11, 12, & 13) had the majority of their seizures during sleep which includes periods of brief arousal (Figure 5A). When we calculated the average daily rate of seizures per sleep stage, we found that for most patients seizures occurred most frequently in NREM1 sleep (patients 2, 9, 10, 11 & 15; Figure 5B). Though the effect of sleep-wake category on the seizure rate was significant for five patients, post-hoc analyses revealed no consistent trends across the cohort (Supplementary Table 1). There was no significant effect of sleep-wake category on seizure duration (Figure 5C; Supplementary Table 2).

**Figure 5.**
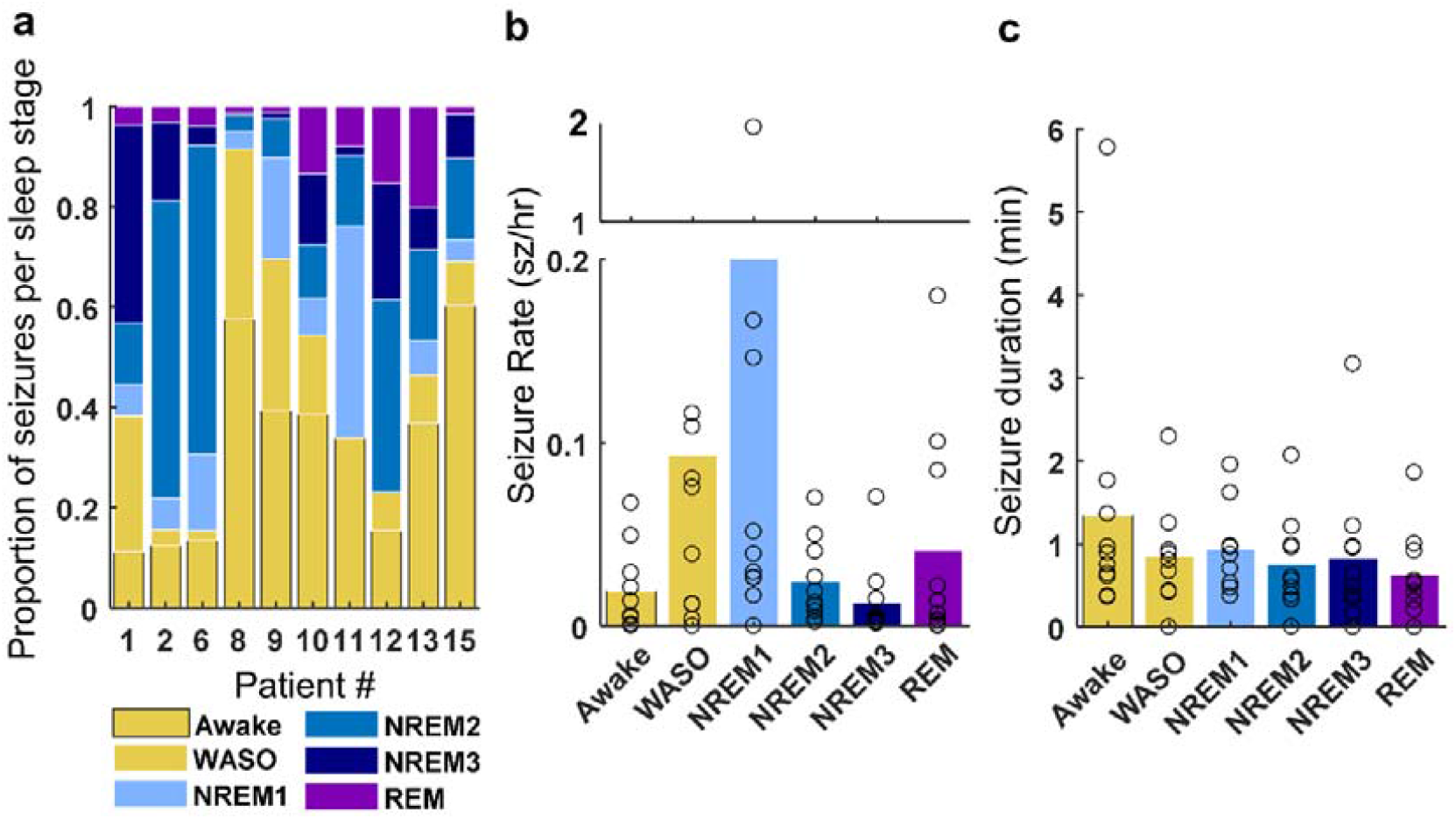
Seizure rate and duration for each sleep-wake state. A) The proportion of seizures occurring in each sleep-wake state are shown for each patient. The group average B) hourly seizure rate (sz/hr) and C) seizure duration (min) is shown for each sleep-wake state. The open circles represent the average for each patient.

### The relation between seizure probability and the day-to-day variability in sleep duration and composition

To assess the interaction between day-to-day deviations in sleep duration and quality and seizure probability, we used logistic regression with random subject-specific intercept. A separate model was used to assess the influence of total sleep duration and the relative proportion of each sleep-wake state, calculated within a 24-hour period. For each patient, days were categorised into decreased, baseline, and increased sleep duration. Baseline sleep was defined as sleep durations falling within the interquartile range (≥25th percentile and ≤75th percentile). Durations below the 25th percentile and above the 75th percentile were classed as decreased and increased sleep duration, respectively. The same method was applied to the relative proportion of each sleep-wake state. The interquartile range for total sleep duration and the proportion of each sleep-wake state is shown in Figure 6A and Figure 7A. Days when a seizure occurred were excluded from these categories. Alteration of sleep duration or sleep-wake state proportion was assumed to play a role if a seizure occurred within 48 hours after an increase or decrease in sleep duration or sleep-wake state proportion. Each patient contributed multiple days of observation into each model, therefore the random effect was the patient to take within subject correlation into account. The sleep categories (decreased, baseline, and increased) were the fixed effects. When age, sex and epileptogenic zone were included in our models we found no significant effect nor any significant confounding influence on the effects of sleep on seizure occurrence (Supplementary Table 3), the variables were therefore excluded from the models.

**Figure 6.**
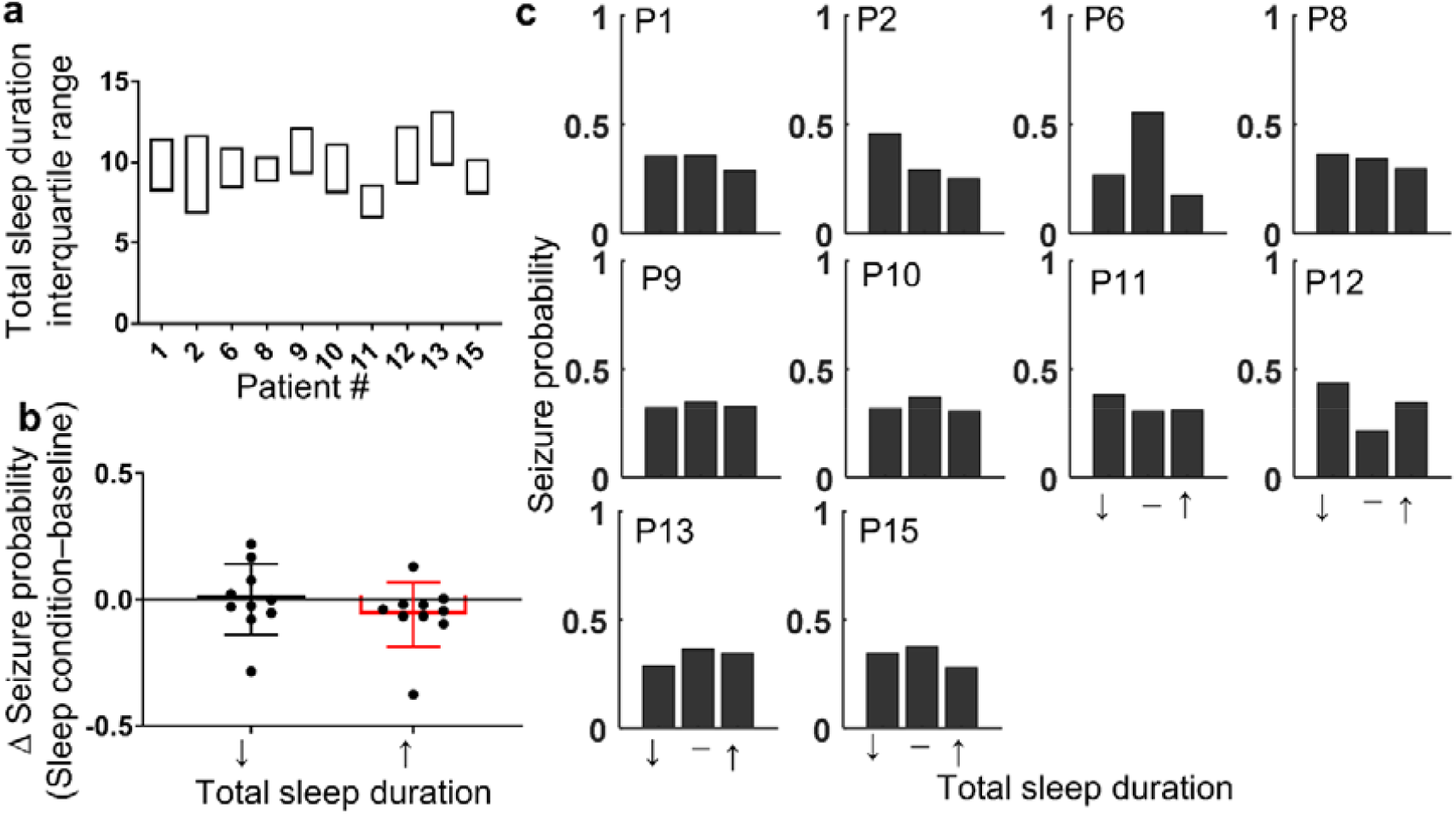
Seizure probability is reduced following a night of increased sleep duration. A) The interquartile range of total sleep duration, calculated in 24-hour periods and including both night sleep and naps, is shown for each patient. B) The population average change in the probability of a seizure occurring in the 48 hours after a night of decreased (↓<25^th^ percentile of total sleep duration) or increased (↑ >75^th^ percentile of total sleep duration) total sleep duration relative to baseline is represented by the hollow bars. The change in probability for individual patients is represented by filled circles. Red bars indicate significant group-level effects. Error bars represent standard deviation. C) For each patient, the normalised probability of a seizure occurring in the 48 hours after a night of decreased (↓), baseline (-), or increased (↑) sleep duration is represented by the filled bars.

**Figure 7.**
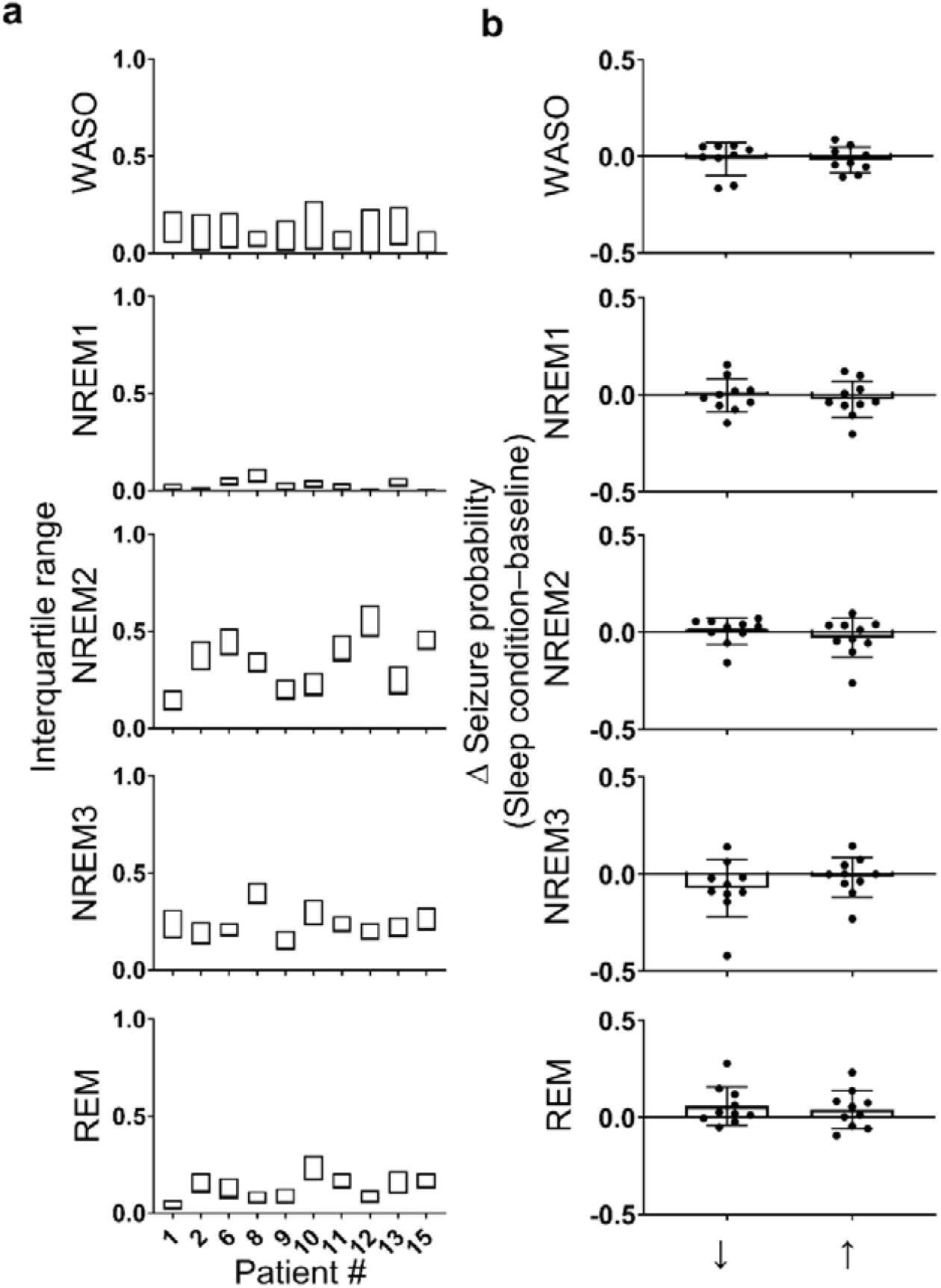
Seizure probability does not change with day-to-day variability in sleep composition. A) The interquartile range for the proportion of each sleep-wake state, calculated in 24-hour periods as a proportion of the total sleep duration, is shown for each patient. B) The population average change in the probability of a seizure relative to baseline is shown for decreased and increased proportions of WASO, NREM1, NREM2, NREM3, and REM sleep by the hollow bars. Individual patient values are indicated by the filled circles. Red bars indicate significant group-level effects. Error bars represent standard deviation.

### Sleep duration

For all but one patient (pt 12), we found that sleeping for longer durations was correlated with a reduced seizure likelihood in the following 48 hours (Figure 6B). Of note, patient 12, who did not follow this trend, experienced the smallest number of seizures (n=13). At the group level, our logistic regression model revealed that when patients slept longer than the 75^th^ percentile there was a 27% reduction in the odds of a seizure in the following 48 hours when compared to baseline (p<0·009; Table 2). For our cohort, the 75^th^ percentile equates to an average of 11·2 (±1·3) hours of sleep, which is 1·66 (±0.52) hours longer than their median sleep duration. Interestingly, sleeping less than the 25^th^ percentile (8·29 ±0·99 hours), which is 1·13 (±0·48) hours less than their median sleep duration, did not have any significant group effect on the odds of a seizure (p=0·48; Table 2). However, using random intercepts modelling, the random effect contributed to the overall model variance significantly, suggesting that subjects responded differently. For four patients (2, 8, 11, & 12), a reduction in total sleep duration was followed by an increased probability of seizure relative to baseline, while for the remaining six patients (1, 6, 9, 13, & 15), the opposite was true (Figure 6B & C). The differences between means for baseline and more/less sleep and the 95% confidence intervals are depicted in Supplementary Figure 2A as bootstrap sampling distributions. There were no consistent trends in the age, sex, epileptogenic zone or medication with regards to the interaction between sleep duration and seizure probability.

**Table 2.** Population level logistic regression model results investigating the interaction between seizure risk and day-to-day variation in sleep duration and sleep composition. A separate model was used for sleep duration and each sleep category proportion.

| Sleep Duration/<br>Composition | Change in<br>sleep relative to<br>baseline | Risk of seizure<br>Odds ratio relative to<br>baseline (95% Confidence<br>interval) | *p*-value<br>(Bonferroni<br>corrected<br>$\alpha=0.025$) | Change in the risk<br>of seizure relative to<br>baseline |
| --- | --- | --- | --- | --- |
| Total Sleep Duration | ↓ | 0.92 (0.74, 1.15) | 0.48 | -- |
|  | ↑ | 0.73 (0.58, 0.92) | **0.009** | ↓ |
| Proportion WASO | ↓ | 1.02 (0.81, 1.27) | 0.89 | -- |
|  | ↑ | 0.86 (0.69, 1.09) | 0.21 | -- |
| Proportion NREM1 | ↓ | 0.91 (0.73, 1.14) | 0.43 | -- |
|  | ↑ | 0.98 (0.78, 1.22) | 0.83 | -- |
| Proportion NREM2 | ↓ | 1.04 (0.84, 1.31) | 0.701 | -- |
|  | ↑ | 1.01 (0.81, 1.27) | 0.93 | -- |
| Proportion NREM3 | ↓ | 0.79 (0.63, 1.001) | 0.052 | -- |
|  | ↑ | 0.92 (0.73, 1.15) | 0.46 | -- |
| Proportion REM | ↓ | 1.29 (1.03, 1.63) | 0.026 | -- |
|  | ↑ | 1.27 (1.02, 1.58) | 0.035 | -- |

### Sleep composition

We anticipated that the odds of seizure might increase following poor-quality sleep, consisting of a higher WASO or NREM1 proportion or a lower NREM3 or REM proportion. However, this was not the case. Instead, any deviation in the proportion of REM sleep from baseline tended to increase the odds of a seizure. A reduced proportion of REM sleep, below the 25th percentile (0·09 ± 0·05) was followed by a 29% increase in the odds of a seizure compared to baseline while an increased proportion of REM sleep (p=0·026), above the 75th percentile (0·17 ± 0·07) was followed by a 27% increase in the odds of a seizure (p=0·035;Table 2). We also observed a lower seizure probability tended to follow a night of reduced NREM3 (p=0·052). However, none of these trends were found to be significant after Bonferroni corrections were applied for multiple comparisons. In general, changes in each sleep type had variable effects across the group, as indicated by the spread of both positive and negative changes in seizure probability relative to baseline (Figure 7B). The differences between means for baseline and more/less REM sleep and the 95% confidence intervals are depicted in Supplementary Figure 2B as bootstrap sampling distributions.

### Sleep duration and composition following seizure occurrence

To characterise sleep duration and composition following a seizure, we divided the data into three categories: days (24-hour periods) that were seizure free (control), days where seizures occurred while the patient was awake (wake sz), and days where seizures occurred during sleep (sleep sz). If a seizure occurred both while awake and asleep, the day was excluded from analysis. The total duration of sleep and the relative proportion of each sleep-wake state were compared across control, wake sz, and sleep sz categories. In general, we found that patients spent more time asleep following a seizure, particularly if the seizure occurred during sleep. However, the composition of sleep was only disrupted if the seizure occurred during sleep.

When compared to control, sleep duration and composition was altered following a seizure (Figure 8A). The occurrence of a seizure correlated with a significant increase in sleep duration, which was most pronounced if the seizure occurred during sleep (Figure 8B; p_Duration_=0·0035, F_1_· _36,12_·_24_=11·16; one-way ANOVA; p_wakeSz_=0.019, p_sleepSz_=0.0045; Dunnett’s multiple comparisons). There were no significant changes in the proportion of each sleep category following a seizure that occurred while awake (Fig. 8C). However, the proportion of time spent awake (in brief arousal during sleep) increased and the time spent in REM sleep decreased when a seizure occurred during sleep, which may suggest that the quality of sleep is reduced by seizure occurrence (Figure 8 C; ANOVA: p_Duration_=0·0035, F_1_·_36,12_· _24_ =11·16; p_WASO_=0·011, F_1_·_69,15_· _24_ =6·531; p_REM_=0·019, F_1_· _49,13_·_44_ =5·999; Dunnett’s multiple comparisons: p_WASO_=0·005, p_REM_=0·0091 (;). We also observed that the proportion of time spent in NREM3 sleep also tended to decrease following a sleep seizure; however, this was not found to be significant (Figure 8C; p_NREM3_=0·090, F_1_·_40,12_· _65_ = 3·161; one-way ANOVA). There were no consistent changes in the proportion of time spent in stage one or two NREM sleep following a seizure (Figure 8C; p_NREM1_=0·20, F_1_· _44,13_·_99_=1·796; p_NREM2_=0·85, F_1_·_86,16_· _78_ =0·1486; one-way ANOVA).

**Figure 8.**
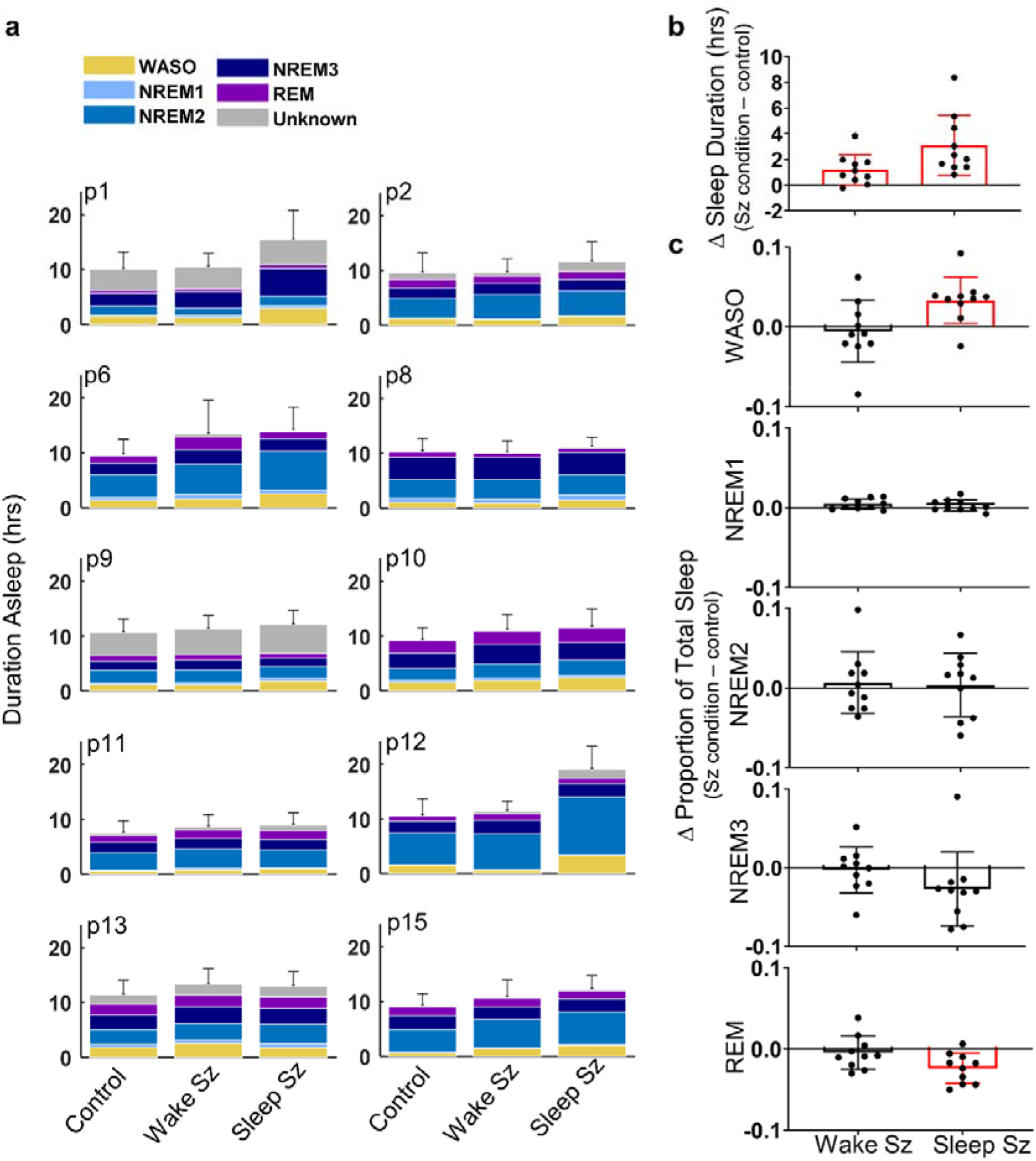
Sleep duration and composition is altered following a seizure. A) For each patient, the average duration asleep (hrs) is illustrated for seizure free (Control), wakeful seizure (Wake Sz), and sleeping seizure (Sleep Sz) categories. B) The average change in sleep duration (hrs) and C) The average change in the proportion of time spent in each sleep-wake state is illustrated for the population (bar) and individual patients (filled circles). Red bars indicate significant group-level effects. Error bars represent standard deviation.

## Discussion

This is the first long-term EEG study to explore the bi-directional relationship between sleep and seizures. In our study, data were collected via intracranial EEG from patients undergoing ordinary life activities. All existing EEG investigations looking at the relationship between seizures and sleep have used data that were collected in hospital settings and of significantly shorter duration, which is unlikely to provide a true representation of relationships that exist between sleep and seizures. We show for the first time that seizure probability changes with day-to-day variations in sleep duration but not sleep composition. An increase in sleep duration was associated with a reduced seizure propensity in the following 48 hours, suggestive of a protective effect. However, counter to expectations, a reduced sleep duration did not consistently increase seizure probability, with grouped effects observed. This may indicate that clinical help for sleep disturbance and seizure problems might benefit from ‘grouped’ attention, as opposed to ‘individual’ or ‘blanket’ treatment methods. Interestingly, improved sleep quality was not associated with reduced seizure risk; no sleep state was found to be significantly associated with seizure likelihood. Finally, we found the occurrence of a seizure correlated with patients sleeping for longer durations and, if the seizure occurred during sleep, the quality of sleep was reduced.

Ours is the first study to use EEG to investigate the interaction between day-to-day deviations in sleep duration and seizure occurrence. We show that an increase in sleep duration by an average of 1·66 ±0·52 or more was associated with a 27% reduction in the odds of seizure in the following 48 hours. While our study design cannot exclude the possibility of reverse causality, where sleep prior to seizures is altered because of pre-seizure effects, our results could suggest that increasing sleep duration is beneficial for patients with epilepsy, reducing seizure risk. Interestingly, for most patients a reduced sleep duration, of 1·13 ±0·48 hours, did not indicate a facilitative role in the generation of seizures. Our mixed effects model indicated that the random effect contributed to the overall model variance significantly, suggesting that subjects respond differently. For four patients, seizure likelihood was higher following a night of reduced sleep duration while for the remaining six patients, seizure likelihood was lower. From a clinical perspective this is a potentially important finding. Indeed, it may suggest that clinical help in this area might benefit from ‘grouped’ attention, as opposed to either, ‘individual’ or ‘blanket’ treatments as the preferred method of addressing sleep disturbance and seizure problems. Larger reports could do well to reflect on clustering mechanisms of treatment administrations.

Although insufficient sleep has long been accepted as a seizure precipitant, supporting evidence predominantly involves profound sleep deprivation (>24 hours).^28^ The influence of small changes in sleep duration (0.5-2 hours) have been scarcely addressed: To our knowledge, there are only four seizure diary studies and they have produced contradictory results.^28^ We believe a large part of this discrepancy may be because of the unreliability of self-reported sleep and seizure diaries with patients reporting less than 50% of their seizures.^17^ Using reliable long-term information about the occurrence of sleep and seizures, our results suggest that mild sleep loss may not be problematic for all patients with epilepsy, though sleeping for longer is beneficial in reducing seizure risk. It is possible that we did not observe a consistent precipitating effect of mild sleep loss because the effect is cumulative and not readily detected by our analytical methods. Chronic partial sleep deprivation occurs more frequently than profound sleep deprivation (>24 hours) yet its effects have not been examined as thoroughly, presumably because they are inherently more difficult to assess.^28^ On average our patient cohort slept for 2.5 hours longer than healthy adults.^26^ This could suggest our patient cohort requires more sleep than healthy adults due to their seizure and/or antiepileptic drug burden. Ultimately, we do not know how much sleep patients with refractory epilepsy require. Thus, it is possible that the apparent neuroprotective effects observed following an increase in sleep duration could reflect recovery from accumulated sleep debt. Alternatively, it could be that the impacts of mild sleep loss are not readily observed in our patient group because the patients were sleeping longer than necessary and thus a small reduction in sleep duration did not exit healthy limits.

Previous research has shown that patients with epilepsy commonly have circadian and infradian rhythms in seizure occurrence.^25,29^ In our patient group both circadian and circaseptan seizure cycles have been described but the relation of these cycles to sleep is yet to be determined.^25^ Here we have shown that, for two patients, seizures tend to occur at the same times of day as their regular sleep onset and offset times. This suggests that sleep timing in particular, could be important in regulating the circadian seizure cycles. We did not observe any clear patterns between sleep timing and seizure occurrence for the circaseptan cycles, though, other sleep factors may contribute more heavily. Further investigation, exploring more cycle durations and aspects of sleep is necessary to fully establish the possible interactions between seizure and sleep-wake cycles.

The effects of day-to-day variations in sleep quality on seizure risk have never before been described. We hypothesised that a higher quality sleep would exert a protective effect. However, we did not find any type of sleep to be significantly associated with seizure propensity. Animal and human studies have indicated that REM sleep, in particular, might be important for people with epilepsy where increasing the proportion of REM sleep may reduce cortical excitability and, thus, seizure risk.^14,15^ Our results do not support this notion, instead we observed trends where both increases and decreases in the proportion of REM sleep were associated with higher odds of a seizure in the following 48 hours. Though these trends were not found to be significant, they would instead imply that unusually high or low REM sleep proportions is problematic for people with epilepsy and thus that regularity of the REM sleep pattern could be more important for seizure management.

Previous studies have indicated the occurrence of a seizure can reduce sleep duration and quality, namely lowering the proportion of REM sleep.^5,16,30,31^ However, these investigations have been conducted in a hospital setting, which is known to alter regular sleep patterns and, therefore, may produce misleading results.^32^ Our data was collected from patients undergoing ordinary life activities. Unlike previous indications, we found that sleep duration was increased following a seizure. This could play an important protective role, helping to reduce the occurrence of subsequent seizures. While we did not observe any significant disruption to the composition of sleep following a wakeful seizure, we found that sleep that was interrupted by a seizure was of poorer quality, including a greater proportion of time spent aroused from sleep and a reduction in REM sleep. This could imply that seizures occur more readily during abnormal sleep or alternatively that sleep is disrupted because of seizure occurrence.

In our investigation, it is important to acknowledge that, while we have months to years of data per patient, our patient cohort was relatively small and highly selected, only including adults with refractory epilepsy, a relatively high seizure burden and a lateralised epileptogenic zone. Thus, the results of our investigation may not apply to all epilepsies in a general way. That said, current information has been based on data with different but significant limitations: Most EEG investigations have used datasets with very few days per patient collected within a hospital environment and often accompanied by medication withdrawal. Thus, we speculate that for patients with refractory epilepsy our results likely provide a more realistic representation of sleep trends. We also acknowledge that the size of our patient cohort may not allow for meaningful exploration of the role of secondary risk factors such as age, sex and epileptogenic zone and their potential confounding influence on the relationship between sleep and seizures. However, when we included these variables into our regression models we did not observe any significant effect or confounding influence, which is in agreement with the existing seizure diary studies.^8,10^ In addition, we did not have access to gold standard sleep information; our sleep-wake scoring was solely based on intracranial EEG. That said, it is not realistic to obtain years of continuous sleep information using polysomnography. The automated methods used in our investigation have previously been shown to produce a high classification accuracy of 94% for awake and stages 2 and 3 NREM sleep.^22^ Furthermore, using both dog and human data we have confirmed our methods perform well for REM sleep with an accuracy of 96% and 95%, respectively (not shown). Altogether, we show that all sleep categories revealed consistent sleep patterns with the expected sleep architecture (Fig. 3-4).”

In this investigation, we did not have information about alcohol consumption, stress, medication compliance, or other similar factors that can influence sleep and seizure occurrence. Thus, it is possible that our results were confounded by other seizure promoting factors. A combined long-term diary and EEG study could help to disentangle the role of sleep from other influences while maintaining the necessary accuracy of sleep and seizure information. However, even if changes in sleep duration or quality are indirectly related to changes in seizure occurrence, our results indicate that sleep provides information that is useful for seizure management and prediction.

## Data Availability

Deidentified recordings of the seizures and some segments of the data used in this study are publicly available on epilepsyecosystem.org. Other relevant data may be made available upon reasonable request by contacting the authors.

## Funding

This research was supported by Australian National Health and Medical Research Council Project Grant 1130468, US National Institutes of Health Grant R01 NS09288203, Czech Technical University in Prague Grant OHK4-026/21 and Epilepsy Foundation of America Innovation Institute, My Seizure Gauge.

## Contributors

Conceived and designed the work: KD, DP, VK, DF, LK, MC, BB, GW. Performed the analysis: KD, DP, VK, VG, PN,GW, LL, RB, FM. Wrote the original draft: KD. Contributed to writing review and editing: KD DP, VK, MM, LK, MC, AB, DG, BB, WD.

## Declaration of Interests

KD, LK and DG report grant support from the Australian National Health and Medical Research Council. VK, BB and GW reports grant support from the USA National Institutes of Health. VK reports institutional support from Czech Technical University in Prague. BB reports grant support from the Epilepsy Foundation of America, equity in Cadence Neuroscience Inc. GW and BB report patents issued and pending in the field of epilepsy and device supplementation for research purposes by Medtronic Inc. WD reports grant support from Eisai Australia and UCB Pharma Global, travel support from UCB Australia, speaker honoraria from Eisai Australia and UCB Pharma Australia, scientific advisory board honoraria from UCB Pharma Australia, Elsai Australia, Liva Nova, and Tilray as well as equity interest in EpiMinder and KeyLead Health. MC reports the position of chair of the SEER Medical board and equity in EpiMinder and SEER Medical. GW reports royalties and equity in NeuroOne Inc., speaker honoraria from Medtronic Inc. and has licenced intellectual property to NeuroOne Inc. and Cadence Neuroscience Inc. All other authors have no conflicts to report.

## Acknowledgements

IEEGPORTAL and Stata 16.0.

## Data sharing

Deidentified recordings of the seizures and some segments of the data used in this study are publicly available on epilepsyecosystem.org. Relevant data that supports the findings of this study and the sleep classifier for each patient can be made available upon reasonable request by contacting the authors.

## Supplementary Material

**Supplementary Table 1.**
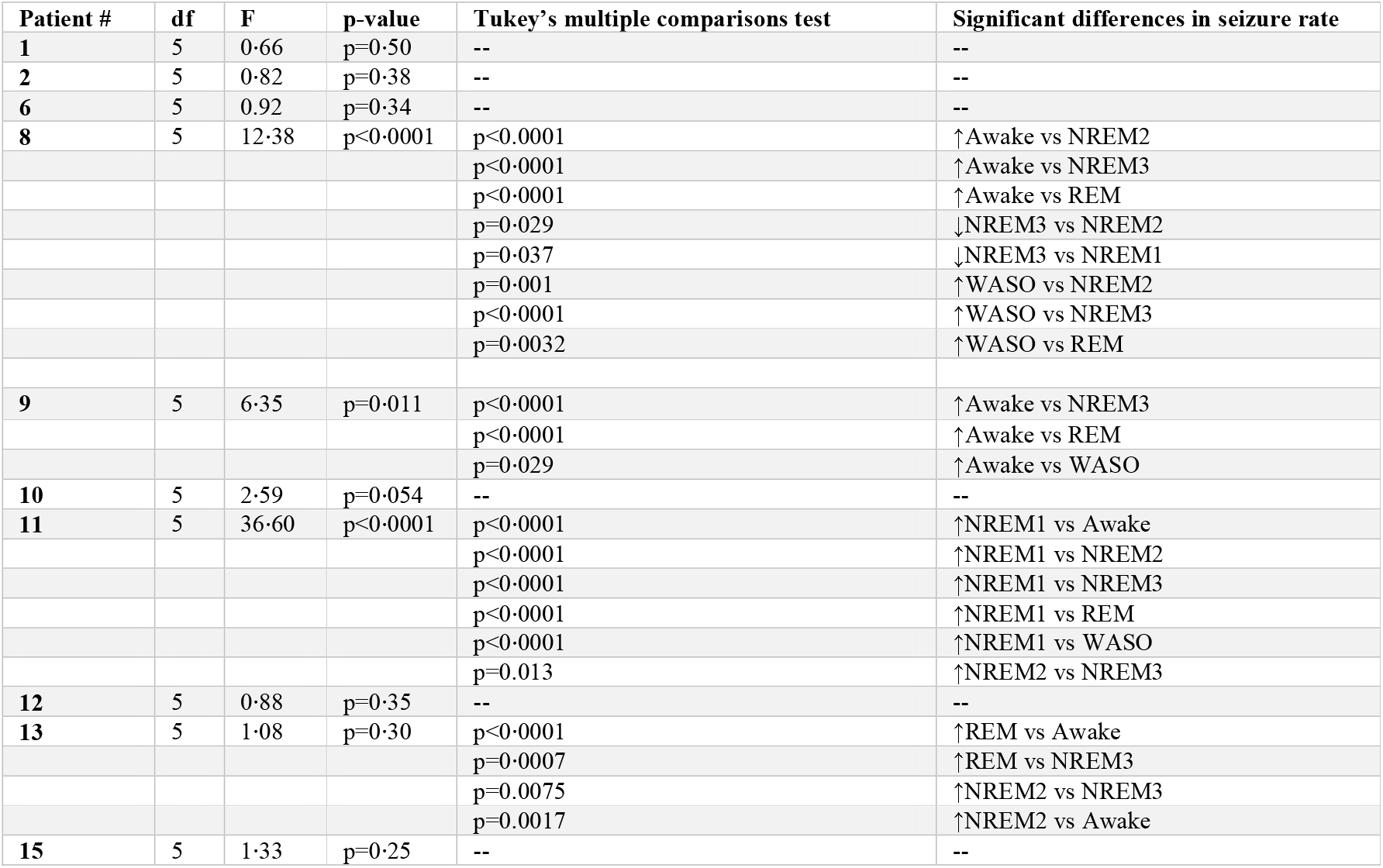
Results of repeated measures one-way ANOVAs looking at the effects of current sleep-wake state on seizure rate.

| Patient # | df | F | p-value | Tukey's multiple comparisons test | Significant differences in seizure rate |
| --- | --- | --- | --- | --- | --- |
| 1 | 5 | 0.66 | p=0.50 | -- | -- |
| 2 | 5 | 0.82 | p=0.38 | -- | -- |
| 6 | 5 | 0.92 | p=0.34 | -- | -- |
| 8 | 5 | 12.38 | p<0.0001 | p<0.0001 | ↑Awake vs NREM2 |
|  |  |  |  | p<0.0001 | ↑Awake vs NREM3 |
|  |  |  |  | p<0.0001 | ↑Awake vs REM |
|  |  |  |  | p=0.029 | ↓NREM3 vs NREM2 |
|  |  |  |  | p=0.037 | ↓NREM3 vs NREM1 |
|  |  |  |  | p=0.001 | ↑WASO vs NREM2 |
|  |  |  |  | p<0.0001 | ↑WASO vs NREM3 |
|  |  |  |  | p=0.0032 | ↑WASO vs REM |
| 9 | 5 | 6.35 | p=0.011 | p<0.0001 | ↑Awake vs NREM3 |
|  |  |  |  | p<0.0001 | ↑Awake vs REM |
|  |  |  |  | p=0.029 | ↑Awake vs WASO |
| 10 | 5 | 2.59 | p=0.054 | -- | -- |
| 11 | 5 | 36.60 | p<0.0001 | p<0.0001 | ↑NREM1 vs Awake |
|  |  |  |  | p<0.0001 | ↑NREM1 vs NREM2 |
|  |  |  |  | p<0.0001 | ↑NREM1 vs NREM3 |
|  |  |  |  | p<0.0001 | ↑NREM1 vs REM |
|  |  |  |  | p<0.0001 | ↑NREM1 vs WASO |
|  |  |  |  | p=0.013 | ↑NREM2 vs NREM3 |
| 12 | 5 | 0.88 | p=0.35 | -- | -- |
| 13 | 5 | 1.08 | p=0.30 | p<0.0001 | ↑REM vs Awake |
|  |  |  |  | p=0.0007 | ↑REM vs NREM3 |
|  |  |  |  | p=0.0075 | ↑NREM2 vs NREM3 |
|  |  |  |  | p=0.0017 | ↑NREM2 vs Awake |
| 15 | 5 | 1.33 | p=0.25 | -- | -- |

**Supplementary Table 2.**
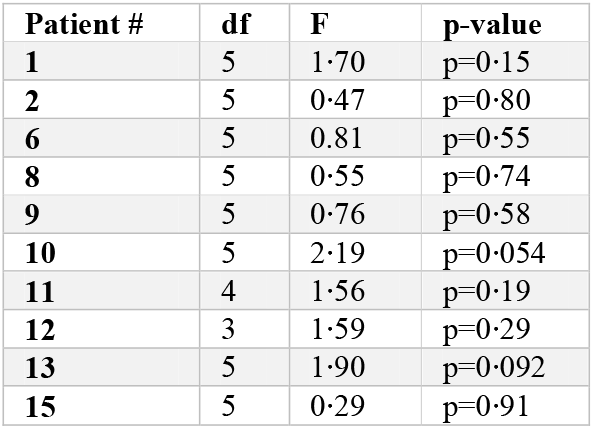
Results of one-way ANOVAs looking at the effects of current sleep-wake on seizure duration.

**Supplementary Table 3.** Results of mixed effects Logistic Regression models

| **Logistic Model** | **Risk Factor** | **Risk of seizure**<br>Odds ratio relative to<br>baseline (95%<br>Confidence interval) | **p-value**<br>(Bonferroni<br>corrected<br>$\alpha=0.25$) | **Change in the**<br>**risk of seizure**<br>**relative to**<br>**baseline** |
| --- | --- | --- | --- | --- |
| Total Sleep Duration | ↓ sleep | 0.92 (0.74, 1.15) | 0.48 | -- |
|  | ↑ sleep | 0.73 (0.58, 0.93) | **0.01** | ↓ |
|  | Age | 1.01 (0.90, 1.15) | 0.83 | -- |
|  | Gender | 2.81 (0.28, 27.93) | 0.38 | -- |
|  | EZ PT vs OP | 0.84 (0.017, 40.98) | 0.93 | -- |
|  | EZ PT vs T | 0.50 (0.012, 21.57) | 0.72 | -- |
|  | EZ PT vs FT | 1.53 (0.026, 87.34) | 0.84 | -- |
|  | EZ OP vs T | 1.68 (0.16, 17.84) | 0.67 | -- |
|  | EZ OP vs FT | 0.55 (0.059, 5.07) | 0.60 | -- |
|  | EZ T vs FT | 0.33 (0.031, 3.41) | 0.35 | -- |
| Proportion WASO | ↓ WASO | 1.02 (0.81, 1.27) | 0.89 | -- |
|  | ↑ WASO | 0.87 (0.69, 1.09) | 0.22 | -- |
|  | Age | 1.01 (0.90, 1.15) | 0.82 | -- |
|  | Gender | 2.95 (0.30, 28.89) | 0.35 | -- |
|  | EZ PT vs OP | 0.81 (0.17, 38.84) | 0.92 | -- |
|  | EZ PT vs T | 0.49 (0.11, 20.85) | 0.71 | -- |
|  | EZ PT vs FT | 1.45 (0.026, 80.83) | 0.85 | -- |
|  | EZ OP vs T | 1.67 (0.15, 17.27) | 0.68 | -- |
|  | EZ OP vs FT | 0.56 (0.061, 5.13) | 0.61 | -- |
|  | EZ T vs FT | 0.34 (0.033, 3.51) | 0.37 | -- |
| Proportion NREM1 | ↓ NREM1 | 0.91 (0.72, 1.14) | 0.43 | -- |
|  | ↑ NREM1 | 0.97 (0.78, 1.22) | 0.82 | -- |
|  | Age | 1.01 (0.89, 1.15) | 0.82 | -- |
|  | Gender | 2.95 (0.302, 28.95) | 0.35 | -- |
|  | EZ PT vs OP | 0.82 (0.017, 38.99) | 0.92 | -- |
|  | EZ PT vs T | 0.50 (0.012, 20.94) | 0.71 | -- |
|  | EZ PT vs FT | 1.46 (0.026, 81.15) | 0.86 | -- |
|  | EZ OP vs T | 1.65 (0.16, 17.28) | 0.68 | -- |
|  | EZ OP vs FT | 0.56 (0.061, 5.13) | 0.61 | -- |
|  | EZ T vs FT | 0.34 (0.033, 3.51) | 0.37 | -- |
| Proportion NREM2 | ↓ NREM2 | 1.04 (0.84, 1.31) | 0.68 | -- |
|  | ↑ NREM2 | 1.01 (0.80, 1.27) | 0.93 | -- |
|  | Age | 1.01 (0.90, 1.15) | 0.82 | -- |
|  | Gender | 2.97 (0.30, 29.01) | 0.35 | -- |
|  | EZ PT vs OP | 0.82 (0.17, 38.79) | 0.92 | -- |
|  | EZ PT vs T | 0.49 (0.12, 20.82) | 0.71 | -- |
|  | EZ PT vs FT | 1.45 (0.026, 80.60) | 0.86 | -- |
|  | EZ OP vs T | 1.65 (0.16, 17.28) | 0.68 | -- |
|  | EZ OP vs FT | 0.56 (0.061, 5.13) | 0.61 | -- |
|  | EZ T vs FT | 0.34 (0.033, 3.51) | 0.37 | -- |
| Proportion NREM3 | ↓ NREM3 | 0.79 (0.63, 1.00) | 0.050 | -- |
|  | ↑ NREM3 | 0.92 (0.74, 1.15) | 0.45 | -- |
|  | Age | 1.01 (0.90, 1.15) | 0.82 | -- |
|  | Gender | 2.95 (0.30, 28.94) | 0.35 | -- |
|  | EZ PT vs OP | 0.82 (0.017, 39.09) | 0.92 | -- |
|  | EZ PT vs T | 0.49 (0.012, 20.88) | 0.71 | -- |
|  | EZ PT vs FT | 1.46 (0.026, 81.56) | 0.85 | -- |
|  | EZ OP vs T | 1.66 (0.16, 17.28) | 0.67 | -- |
|  | EZ OP vs FT | 0.56 (0.061, 5.13) | 0.61 | -- |
|  | EZ T vs FT | 0.34 (0.033, 3.51) | 0.36 | -- |
| Proportion REM | ↓ REM | 1.29 (1.03, 1.63) | 0.025 | -- |
|  | ↑ REM | 1.26 (1.01, 1.57) | 0.035 | -- |
|  | Age | 1.01 (0.90, 1.14) | 0.82 | -- |
|  | Gender | 2.98 (0.30, 29.35) | 0.35 | -- |
|  | EZ PT vs OP | 0.81 (0.017, 38.95) | 0.92 | -- |
|  | EZ PT vs T | 0.49 (0.02, 20.97) | 0.71 | -- |
|  | EZ PT vs FT | 1.44 (0.026, 80.41) | 0.86 | -- |
|  | EZ OP vs T | 1.64 (0.16, 17.29) | 0.68 | -- |
|  | EZ OP vs FT | 0.57 (0.061, 5.19) | 0.62 | -- |
|  | EZ T vs FT | 0.34 (0.033, 3.56) | 0.37 | -- |

**Supplementary Figure 1.**
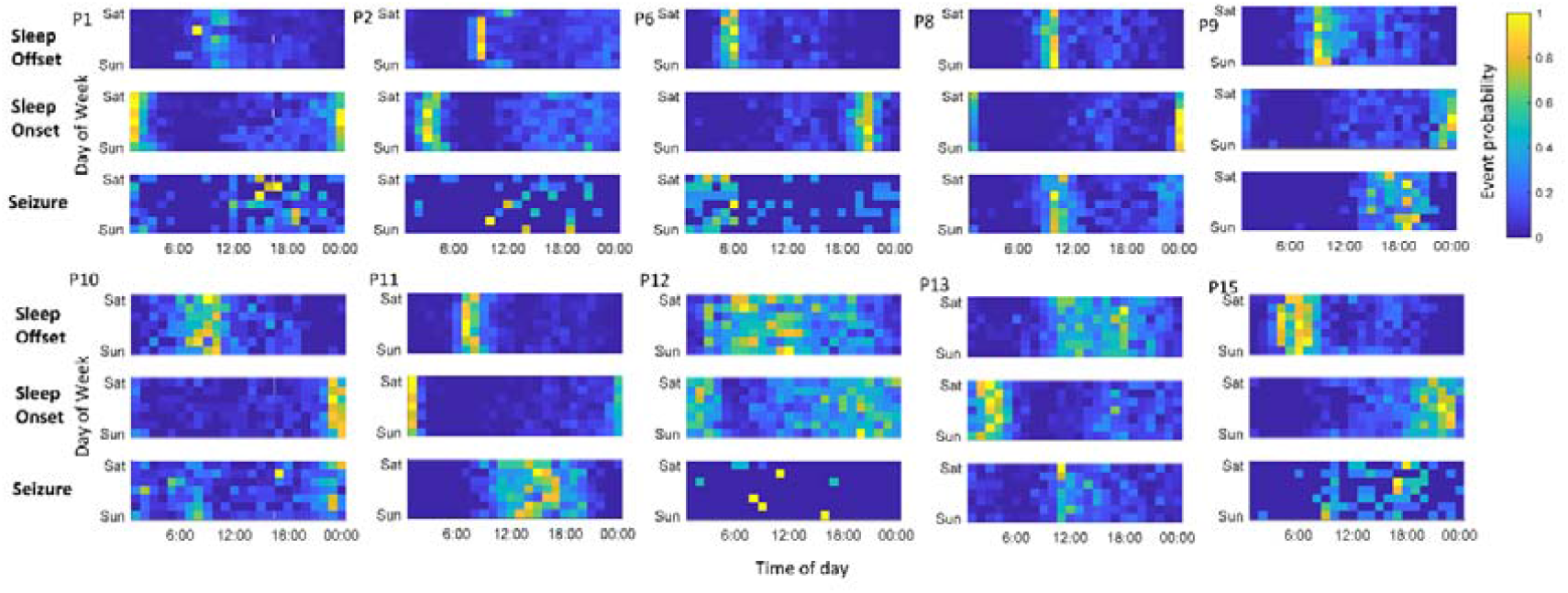
Circaseptan cycle of seizure, sleep onset and sleep offset. The probability of sleep offset (top), sleep onset (middle) and seizure (bottom) are indicated for each patient. Yellow indicates a higher probability of the event and blue a low probability.

**Supplementary Figure 2.**
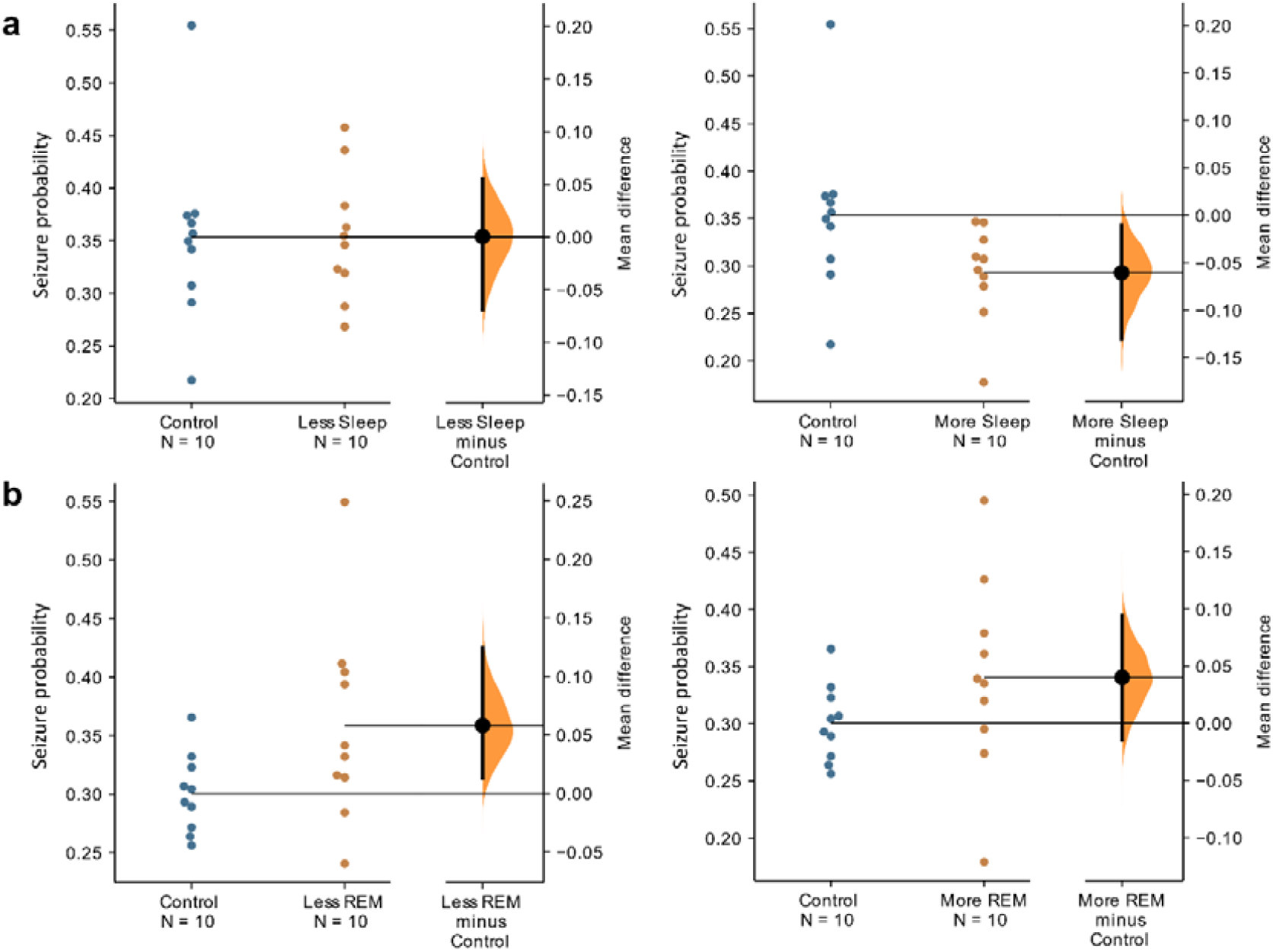
Seizure probability following changes in sleep duration and REM proportion. The Gardner-Altman^1^ estimation plots demonstrate the difference between the means for A (left) control and less sleep, A (right) control and more sleep, B (left) control and less REM and B (right) control and more REM. For each figure the patient seizure probabilities are plotted on the left axes (coloured dots); the mean difference is plotted on a floating axis on the right as a bootstrap sampling distribution (5000 samples; bias corrected and accelerated). The mean difference is depicted as a dot; the 95% confidence interval is indicated by the ends of the vertical error bar.

## Notes

### Author Declarations

Approved by the Human Research Ethics Committees of the three participating clinical centres: Austin Health, The Royal Melbourne Hospital, and St Vincent's Hospital of the Melbourne University Epilepsy Group (LRR145/13).

